# Breastfeeding intentions and breastfeeding reality among first-time mothers living with obesity: A qualitative thematic analysis

**DOI:** 10.64898/2026.08.17.26360590

**Authors:** Doris Keller, Evelyne M. Aubry

**Affiliations:** Department of Health Professions, Bern University of Applied Sciences, Bern, Switzerland

**Keywords:** breastfeeding, obesity, first-time mothers, breastfeeding intention, postpartum care, midwifery care

## Abstract

**Background:** Maternal obesity is associated with lower breastfeeding initiation, shorter breastfeeding duration and lower rates of exclusive breastfeeding. Although breastfeeding intention predicts initiation, little is known about how first-time mothers living with obesity experience the transition from antenatal intention to postpartum reality.

**Aim:** This qualitative study explored breastfeeding expectations, intentions, attitudes and early breastfeeding challenges among first-time mothers living with obesity in Switzerland.

**Methods:** Seven semi-structured interviews were conducted with first-time mothers living with obesity in German-speaking Switzerland. Interviews were audio-recorded, transcribed verbatim and analysed using reflexive thematic analysis according to Braun and Clarke.

**Results:** All participants intended to breastfeed and described breastfeeding as part of motherhood. However, most experienced a postpartum reality that differed from their expectations. Three themes were developed: antenatal engagement with and expectations of breastfeeding, postpartum breastfeeding reality, and everyday breastfeeding life. Women received little antenatal breastfeeding counselling and faced challenges related to medicalised birth, delayed lactogenesis II, breast anatomy, pain, insufficient milk supply, pumping and inconsistent professional support. Several women described feelings of failure when breastfeeding did not work as hoped.

**Conclusion:** First-time mothers living with obesity may have strong breastfeeding intentions but still experience major challenges after birth. Breastfeeding support should start during pregnancy and be realistic, respectful and weight-inclusive.

## Introduction

Obesity is also relevant in pregnancy. In Switzerland, the prevalence of maternal obesity is 7.2%, meaning that more than 6,000 women per year enter pregnancy with obesity (Aubry et al., 2019). Maternal obesity is associated with increased risks for women and infants, including gestational diabetes, hypertensive disorders, small for gestational age, preterm birth, intrauterine fetal death, operative birth, caesarean birth, postpartum infection and thromboembolic events (Aubry et al., 2019; Catalano & Shankar, 2017; Corcoran et al., 2013; DeJoy et al., 2016; Ellis et al., 2019; Fakhraei et al., 2022; Marchi et al., 2015; Maughan et al., 2022; Torloni et al., 2009; Wang et al., 2013).

Because of these risks, pregnant women living with obesity are often treated as a risk group and receive care that is strongly focused on surveillance and intervention. The German S3 guideline on obesity and pregnancy recommends close monitoring, ultrasound examinations, antenatal fetal monitoring, birth in a centre hospital and an intervention-oriented birth management when needed (Arbeitsgemeinschaft der Wissenschaftlichen Medizinischen Fachgesellschaften, 2019). However, risk-focused care may also have negative effects. Women living with obesity may feel objectified when they are not seen as individuals with needs, wishes and experiences (Smith & Lavender, 2011; Thorbjörnsdottir et al., 2020).

Breastfeeding has health benefits for infants and mothers. Breastfed children have lower risks of several acute and chronic conditions, including otitis media, diarrhoeal disease, lower respiratory tract disease, sudden infant death syndrome, childhood leukaemia, diabetes, obesity, asthma and atopic dermatitis (Meek et al., 2022; Stoody et al., 2019). For mothers, breastfeeding is associated with lower risks of type 2 diabetes, breast cancer, ovarian cancer, uterine cancer and hypertension (Feltner et al., 2018; Meek et al., 2022).

Despite these benefits, mothers living with obesity have less favourable breastfeeding outcomes than mothers with normal weight. In industrialised countries, mothers living with obesity have a lower rate of breastfeeding initiation and are less likely to still be breastfeeding six months postpartum (Bever Babendure et al., 2015). Some studies suggest that women with a BMI of 30 kg/m^2^ or higher intend to breastfeed as often as women with normal weight, but breastfeed for a shorter time and are less likely to breastfeed exclusively (Hauff et al., 2014; Jarlenski et al., 2014; Marshall et al., 2018). Other studies describe maternal obesity as a major risk factor for lower breastfeeding initiation, shorter breastfeeding duration and lower exclusivity of breastfeeding (Bever Babendure et al., 2015; Turcksin et al., 2014).

Different factors may influence breastfeeding among mothers living with obesity. Medical and birth-related factors include induction of labour, synthetic oxytocin, epidural analgesia and caesarean birth, which may affect breastfeeding outcomes (Andrew et al., 2022). Biological factors include delayed lactogenesis II and altered hormonal responses. Lactogenesis II refers to the onset of copious milk production after birth. It may be delayed among mothers living with obesity, and a reduced prolactin response to infant sucking may affect milk production (Bever Babendure et al., 2015; Massov, 2015). Practical factors include difficulties with positioning and latching, large breasts, flat nipples, the need to support breast tissue during feeding, and the use of breast pumps or nipple shields (Garner et al., 2017; Massov, 2015). Psychosocial factors include reduced confidence in the body, reduced breastfeeding self-efficacy, limited social support and discomfort with breastfeeding in public (Claesson et al., 2018; Lyons et al., 2018).

Breastfeeding intention is defined as a pregnant woman’s intention to feed her child with breast milk, either directly at the breast or by bottle. It is considered a strong predictor of breastfeeding initiation after birth (Achike & Akpinar-Elci, 2021). Factors that influence breastfeeding intention and duration include maternal age, occupation, having been breastfed as a child, smoking, obesity, social support, birth complications, caesarean birth, anxiety and self-efficacy (Achike & Akpinar-Elci, 2021).

There is a need for qualitative research that explores how mothers living with obesity understand breastfeeding before birth and how they experience breastfeeding after birth. The personal perspective of these women is needed to develop better breastfeeding support.

The aim of this study was to explore breastfeeding expectations, intentions and attitudes among first-time mothers living with obesity. The study also aimed to describe the challenges, barriers and difficulties these women experienced at the beginning of breastfeeding, and to examine how antenatal breastfeeding intentions differed from postpartum breastfeeding reality.

## Materials and methods

### Study design

This study used a qualitative exploratory design with semi-structured interviews. This design was chosen to gain insight into the subjective perspectives of first-time mothers living with obesity. The study focused on antenatal breastfeeding expectations, intentions and attitudes, as well as on factors and experiences around breastfeeding initiation and the postpartum breastfeeding period (Helfferich, 2011b).

A theory-based interview guide was developed using the SPSS principle by Helfferich. SPSS refers to collecting, checking, sorting and subsuming interview questions (Helfferich, 2011a). The guide included questions about antenatal breastfeeding expectations, influencing factors, difficulties around breastfeeding initiation and postpartum breastfeeding reality.

### Participants and sampling

The target group was first-time mothers living with obesity. Inclusion criteria were: preconception BMI of 30 kg/m^2^ or higher, residence in Switzerland, ability to communicate in German or English, birth of a live-born child in 2023, and breastfeeding experience after birth for a shorter or longer period. Women who were still breastfeeding at the time of the interview could also participate.

Women were excluded if they were multiparous, had no breastfeeding experience, had given birth before 2023, did not speak German or English, or lived outside Switzerland.

### Recruitment

Seven women were recruited between September and November 2023. Recruitment took place through flyers, social media channels of Breastfeeding Promotion Switzerland and the professional network of the researcher. The researcher was a self-employed midwife and had previously worked as a hospital midwife in different clinics in German-speaking Switzerland.

When a midwife knew a woman who met the inclusion criteria, she asked for permission to share her contact details with the researcher. The researcher then contacted the woman by telephone, answered questions and arranged an interview. Written informed consent was obtained before the interview.

### Data collection

Each interview started with an open narrative prompt. This was followed by semi-structured questions related to the research questions (Helfferich, 2011a).

Seven semi-structured interviews were conducted between 2 November and 21 December 2023. All participants had given birth to a live-born child in 2023. One participant had given birth to twins. With one exception, the interviews took place in the participants’ homes, often in the presence of their children. The interviews lasted between 36 and 60 minutes, with a mean duration of 53 minutes.

### Data analysis

The interviews were audio-recorded and transcribed verbatim. The simple transcription rules by Dresing and Pehl were used (Dresing & Pehl, 2015).

The data were analysed using thematic analysis according to Braun and Clarke. Thematic analysis is a method for identifying, analysing and developing themes from qualitative datasets. It is flexible and not tied to one theoretical framework (Boyatzis, 1998; Braun & Clarke, 2006).

The analysis followed six steps. First, the researcher became familiar with the full dataset by listening to the audio files several times, reading the transcripts and writing first ideas in a personal journal. Second, initial codes were generated. Two transcripts were coded independently by the researcher and two additional researchers. These codes were discussed and triangulated. The remaining five transcripts were then coded with new inductive codes and with already established codes. Third, codes were grouped into potential themes. Fourth, themes were reviewed and refined. Fifth, the themes were named and defined. Sixth, the themes were written into a report and illustrated with data extracts (Braun & Clarke, 2022). The analysis was conducted using MAXQDA 2022.

### Ethical considerations

A clarification of responsibility was submitted to the Ethics Committee of the Canton of Bern before the study began. The Ethics Committee reviewed the study protocol under Req-2023-00971 and confirmed on 16 August 2023 that the study did not fall under the Swiss Human Research Act.

All participants gave written informed consent. They were informed about the purpose, content and procedure of the study and about data anonymisation. The anonymised data were stored on a password-protected external hard drive. Participation was voluntary. Participants were informed that they could withdraw their consent at any time without giving a reason.

## Results

### Participant characteristics

Seven first-time mothers living with obesity participated in the study. The women were in avarage 31 years old. All were married. Most lived in rural areas of German-speaking Switzerland and had completed vocational education. Their mean BMI was 36 kg/m^2^. All participants had a positive attitude toward breastfeeding.

One participant had a spontaneous birth. Four participants had a caesarean birth. Three children, including twins, were born preterm. Two participants were able to breastfeed exclusively. The other participants breastfed partially, expressed breast milk or had stopped breastfeeding by the time of the interview.

The thematic analysis generated 194 initial codes, 43 potential themes and three main themes:

1. Antenatal engagement with and expectations of breastfeeding
2. Postpartum breastfeeding reality
3. Everyday breastfeeding life

### Theme 1: Antenatal engagement with and expectations of breastfeeding

All participants wanted to breastfeed after birth. For many women, breastfeeding was not experienced as a decision that needed to be made. It was described as something that had been clear from the beginning. The women saw breastfeeding as part of becoming a mother. They associated it with closeness, bonding and doing something good for their child.

### One participant explained

> “For me it was actually clear: I want to breastfeed the child. It was not really a decision of whether I wanted to breastfeed or not. The decision was more: if it works, then I want to breastfeed.”
>
> T2

The women prepared for breastfeeding in different ways. Some did not actively search for much information and wanted to wait and see how breastfeeding would be for them. Others read books, searched online, listened to podcasts or talked with friends and family members. These different sources sometimes created uncertainty because the women received many opinions and had to decide which information was relevant.

### One participant described this information search as confusing

> “I informed myself in many different places. It became quite a mix of many things.”
>
> T4

The participants had different plans for breastfeeding duration. Some hoped to breastfeed for a longer time. Others planned to breastfeed until they returned to paid work three to four months after birth. Most women knew that breastfeeding could be difficult. They tried not to put too much pressure on themselves and wanted to stay open to what would happen after birth.

Practical preparation was also important. Several women bought breastfeeding aids, such as breastfeeding pillows, breast pumps, bottles and formula. Some followed recommendations from friends or family members. A few participants were advised to express and store colostrum before birth. For some women, antenatal colostrum expression strengthened confidence in their ability to produce breast milk. For others, being unable to express colostrum caused uncertainty.

Partners and the social environment influenced breastfeeding expectations. Most partners understood the benefits of breastfeeding and were supportive without putting pressure on the women. Family members, sisters, cousins and colleagues who had breastfed served as positive examples. Participants also described breastfeeding as socially accepted. However, some noted that breastfeeding in public was not very visible. Some felt breastfeeding was intimate and did not want to expose their large breasts in public.

Health professionals had little influence on antenatal breastfeeding expectations. All participants received antenatal care from gynaecologists. Antenatal midwifery care was the exception. Breastfeeding was not discussed in detail during pregnancy. None of the women received information about possible breastfeeding challenges linked to obesity. In antenatal classes led by midwives, breastfeeding was mentioned only briefly and in general terms. Even women who were hospitalised for a longer time during pregnancy did not receive specific breastfeeding preparation.

### One participant stated

> “During pregnancy, it was never really a topic. Neither with the gynaecologist nor in hospital during check-ups. No one talked about what breastfeeding would look like afterwards.”
>
> T7

The participants placed high expectations on postpartum midwifery support. They expected theoretical and practical help in the early breastfeeding period. For some women, professional support was closely linked to the belief that breastfeeding would be possible if the right support was available.

### Theme 2: Postpartum breastfeeding reality

Only one participant experienced breastfeeding initiation as she had imagined it during pregnancy. For the other women, postpartum breastfeeding reality differed from their expectations. Reasons included preterm birth, caesarean birth, pain, stress, delayed lactogenesis II, breast anatomy and insufficient milk supply.

### One participant who experienced breastfeeding as expected said

> “Actually, it was exactly as I had imagined it. I was still convinced: I can breastfeed. I will breastfeed. And it simply worked.” T5

For most women, the start was different. One participant became the mother of preterm twins at 30 weeks of pregnancy. Her children stayed in neonatal care, and direct breastfeeding was postponed until they had enough strength and had learned the necessary function. Other participants described breastfeeding initiation as painful, stressful and more complex than expected.

### One woman described the pain and stress

> “My nipples hurt so much and were inflamed from all the latching. It did not work with large breasts and nipples that are far down and flat. It was very stressful.”
>
> T7

Birth experience shaped breastfeeding reality. One participant had an uncomplicated spontaneous birth. The other women gave birth by vacuum extraction, planned caesarean birth or unplanned caesarean birth, often after induction of labour. Several participants reflected that birth stress, caesarean birth and delayed milk production influenced breastfeeding. One woman felt that her uncomplicated birth supported breastfeeding because she had enough energy after birth.

Delayed lactogenesis II was common. More than half of the participants reported that several days after birth they had no colostrum or that lactogenesis II had not yet occurred. This caused concern and uncertainty.

### One participant said

> “I had no milk. I was in hospital for one week and I had nothing, not one drop.”
>
> T4

Breast anatomy was another important factor. Large breasts, flat nipples and nipples that were difficult for the infant to latch onto made breastfeeding more difficult. Some women found breastfeeding positions complicated and physically demanding. They needed practical help to find positions that worked for their bodies.

The participants realised that breastfeeding involved many techniques, positions and decisions. This was different from their expectation that breastfeeding would be more intuitive. Some women developed self-doubt and wondered whether they were able to meet the demands of breastfeeding. When breastfeeding did not work as hoped, and formula was needed, some women felt they had failed.

### One participant described this emotional moment

> “That was the only moment when I burst into tears and thought: Am I a bad mother if I cannot breastfeed?”
>
> T3

Five of the seven participants used an electric breast pump. Pumping was used instead of, or in addition to, breastfeeding. For many women, pumping produced little milk and did not solve pain or low milk supply. It added work to an already difficult situation. The combination of breastfeeding, pumping, storing milk and bottle-feeding was experienced as tiring and time-consuming.

Professional support after birth was important, but experiences differed. The women appreciated individual information, practical help, praise and confirmation. They wanted support that helped them help themselves. They also valued honest assessments of breastfeeding and milk supply.

### One participant described how practical support helped her

> “The nurse had time to breastfeed with me, also with the large breasts, and to show me how to position the baby. That helped me to gain some confidence.” T2

At the same time, several participants described insufficient or inconsistent support. Some received contradictory advice from different nurses or midwives. Others felt left alone with pain, worries and uncertainty. The women reported that they had not been prepared for breastfeeding as a process. They also reported that no one explained possible links between higher body weight, delayed lactogenesis II, birth mode and breastfeeding difficulties. The clinical focus was often on milk quantity and infant weight gain, while the women had to find their own way to reach these goals.

Some participants experienced strong pressure to breastfeed on the postnatal ward. Alternative feeding methods, such as combining breast milk and formula, were often not discussed. Some women felt judged when they could not breastfeed or no longer wanted to breastfeed. One participant described a situation in which a nurse handled her breast and pushed the crying infant onto the breast. This was experienced as intrusive and not supportive.

### The participant explained

> “I was sitting there and my arms were hanging down. I thought: I also have to learn this. What am I supposed to do if you take my child, push him onto my breast, squeeze my breast and push it into his mouth?” T4

Overall, participants wanted more respectful, sensitive and practical breastfeeding support.

### Theme 3: Everyday breastfeeding life

After discharge from hospital, the women described everyday life with their infant as demanding. Breastfeeding was time-consuming and was experienced as a never-ending loop. Mothers described their new role as a 24-hour job.

Everyday life required flexibility. The women learned to adjust their expectations and respond to changes. Some described the need to try out different ways of managing daily life with the baby. Unexpected events, such as breastfeeding refusal or mastitis, could still occur even after the first difficulties seemed to have improved.

Women who stopped breastfeeding did not make this decision lightly. Some found it difficult to accept that breastfeeding had not worked as hoped. One woman described feeling like a criminal when she told people that her child was fed only with formula. At the same time, women who had stopped breastfeeding described relief, less stress, better physical well-being and more time for other things. Formula feeding made them more independent because other caregivers could also feed the child. Nevertheless, some women continued to wonder whether they had given up too early or had not been patient enough.

## Discussion

This study explored breastfeeding expectations, intentions and attitudes among first-time mothers living with obesity and compared these with postpartum breastfeeding reality. All participants intended to breastfeed during pregnancy. They described breastfeeding as a natural part of motherhood and breast milk as the gold standard for infant feeding. They also valued breastfeeding because of its practicality and its role in bonding.

This finding differs from studies suggesting that women living with obesity are less likely than women with normal weight to intend to breastfeed (Achike & Akpinar-Elci, 2021; Amir & Donath, 2007). In the present study, breastfeeding intention was strong. However, for most participants, postpartum breastfeeding reality did not match antenatal expectations. Most women could not breastfeed exclusively, and only one participant experienced breastfeeding initiation as expected.

One possible explanation is the influence of birth-related factors. Almost all participants experienced an intervention-rich and medicalised birth. This aligns with research showing that birth care for women living with obesity is often medically oriented and focused on risk (Kerrigan et al., 2015). Interventions such as synthetic oxytocin, epidural analgesia and caesarean birth may affect breastfeeding outcomes (Andrew et al., 2022). Epidural opioids may influence infant feeding reflexes, and synthetic oxytocin may affect oxytocin receptors in the breast and the milk ejection reflex (Beilin et al., 2005; Odent, 2013). Skin-to-skin contact, undisturbed bonding and breastfeeding in the first hour after birth can support breastfeeding, including after caesarean birth when the clinical situation allows it (Takahashi et al., 2017; Ulfa et al., 2023).

Delayed lactogenesis II and anatomical challenges were also important. More than half of the participants described delayed milk production. Several women also experienced difficulties with positioning and latching due to large breasts, flat nipples or other anatomical factors. These findings are consistent with earlier research describing additional breast tissue, large breasts and unfavourable areola or nipple anatomy as possible barriers to breastfeeding among mothers living with obesity (Bever Babendure et al., 2015; Garner et al., 2014; Keely et al., 2015).

The findings also show a gap between strong antenatal intention and limited antenatal preparation. Breastfeeding was not addressed in detail during pregnancy. The participants did not receive specific information about breastfeeding with obesity or about possible challenges such as delayed lactogenesis II, positioning problems or the influence of birth mode. This is important because mothers living with obesity may need additional and longer breastfeeding support to initiate and maintain breastfeeding (Achike & Akpinar-Elci, 2021; Garner et al., 2017; Lyons et al., 2018).

The lack of preparation and the difficult postpartum reality contributed to feelings of insufficiency and failure. When breastfeeding did not work and formula was needed, some women felt that they had failed. Brown (2016) described that such stress can negatively influence breastfeeding initiation. A focus on partial breastfeeding and step-by-step goals may reduce pressure and support maternal well-being (Brown, 2016). Lyons et al. (2023) also suggested that mothers living with obesity may benefit from planning breastfeeding for short periods and then reviewing and extending these goals.

The findings suggest that breastfeeding support for mothers living with obesity should be realistic, individual and weight-inclusive. It should prepare women for possible challenges without increasing fear or shame. It should also avoid reinforcing weight stigma. This is important because health professionals may feel uncomfortable discussing weight, fear offending women or lack the knowledge and skills to discuss obesity effectively (Dieterich & Demirci, 2020; Lindberg et al., 2023).

In the present study, weight and breastfeeding-related challenges linked to obesity were not discussed during antenatal care. This silence may have left women without the information they needed. Antenatal breastfeeding care should therefore include tailored counselling about breastfeeding with obesity. This could include information about lactogenesis II, birth mode, positioning, breast anatomy, expressing milk, partial breastfeeding and combined feeding. Counselling should also support maternal autonomy and reduce guilt.

Effective breastfeeding interventions for mothers living with obesity should promote continuous midwifery care across pregnancy, birth, the postpartum period and breastfeeding (Conrey et al., 2024). Such care may help women to build realistic expectations, recognise possible barriers early and receive support before difficulties become overwhelming.

### Strengths and limitations

A strength of this study is the qualitative design, which allowed a deep exploration of personal views and experiences. The use of semi-structured interviews and thematic analysis was suitable for answering the research questions. Another strength is the professional background of the researcher as a midwife and IBCLC. This helped to build trust and to address sensitive topics. At the same time, this background may also have influenced the research process. The researcher therefore reflected on her assumptions in the research team.

Another strength is that all participants were first-time mothers. Their breastfeeding expectations were based on theoretical knowledge rather than previous personal breastfeeding experience. This allowed the study to compare antenatal expectations with first postpartum breastfeeding experiences. The participants lived in six different cantons of German-speaking Switzerland and had been cared for by different medical and nursing teams.

The study also has limitations. The sample was small. All participants were born, raised and socialised in Switzerland. This limits transferability to other countries, cultural groups and healthcare systems. However, the interviews generated rich data and provide insight into the experiences of first-time mothers living with obesity in German-speaking Switzerland.

## Conclusion

This qualitative study shows that first-time mothers living with obesity may have strong antenatal intentions to breastfeed. However, their postpartum breastfeeding reality can differ greatly from their expectations. In this study, most women experienced difficulties with breastfeeding initiation, delayed lactogenesis II, pain, stress, anatomical challenges, pumping, formula supplementation and inconsistent professional support.

The study also shows that breastfeeding with obesity was not addressed during antenatal care. Participants did not receive tailored information about possible breastfeeding challenges related to higher body weight, birth mode or delayed milk production. This lack of preparation contributed to uncertainty, pressure and feelings of failure when breastfeeding did not work as expected.

Future breastfeeding support for mothers living with obesity should start during pregnancy. It should be realistic, respectful and tailored to women’s needs. Continuous midwifery support across pregnancy, birth, the postpartum period and breastfeeding may help women prepare for possible barriers, strengthen breastfeeding confidence and reduce guilt or shame.

## Data Availability

All data produced in the present study are available upon reasonable request to the authors

## Contribution

DK and EA conceived the study and developed the methodology, DK carried out the investigation, curated and analyzed the data, EA supervised the project, led project administration, DK and EA wrote the original draft

